# Accelerated waning of immunity to SARS-CoV-2 mRNA vaccines in patients with immune mediated inflammatory diseases

**DOI:** 10.1101/2022.01.26.22269856

**Authors:** Roya M. Dayam, Jaclyn C. Law, Rogier L. Goetgebuer, Gary Y. C. Chao, Kento T. Abe, Mitchell Sutton, Naomi Finkelstein, Joanne M. Stempak, Daniel Pereira, David Croitoru, Lily Acheampong, Saima Rizwan, Klaudia Rymaszewski, Raquel Milgrom, Darshini Ganatra, Nathalia V. Batista, Melanie Girard, Irene Lau, Ryan Law, Michelle W. Cheung, Bhavisha Rathod, Julia Kitaygorodsky, Reuben Samson, Queenie Hu, W. Rod Hardy, Nigil Haroon, Robert D. Inman, Vincent Piguet, Vinod Chandran, Mark S. Silverberg, Anne-Claude Gingras, Tania H. Watts

## Abstract

**Background:** Limited information is available on the impact of immunosuppressants on COVID-19 vaccination in patients with immune-mediated inflammatory diseases (IMID).

**Methods:** This observational cohort study examined the immunogenicity of SARS-CoV-2 mRNA vaccines in adult patients with inflammatory bowel disease, rheumatoid arthritis, ankylosing spondylitis, or psoriatic disease, with or without maintenance immunosuppressive therapies. Antibody and T cell responses to SARS-COV-2, including neutralization against SARS-CoV-2 variants were determined before and after 1 and 2 vaccine doses.

**Results:** We prospectively followed 150 subjects, 26 healthy controls, 9 IMID patients on no treatment, 44 on anti-TNF, 16 on anti-TNF with methotrexate/azathioprine (MTX/AZA), 10 on anti-IL-23, 28 on anti-IL-12/23, 9 on anti-IL-17, and 8 on MTX/AZA. Antibody and T cell responses to SARS-CoV-2 were detected in all participants, increasing from dose 1 to dose 2 and declining 3 months later, with greater attrition in IMID patients compared to healthy controls. Antibody levels and neutralization efficacy against variants of concern were substantially lower in anti-TNF treated patients than in healthy controls and were undetectable against Omicron by 3 months after dose 2.

**Conclusions:** Our findings support the need for a third dose of mRNA vaccine and for continued monitoring of immunity in these patient groups.

**Funding:** Funded by a donation from Juan and Stefania Speck and by Canadian Institutes of Health (CIHR) /COVID-Immunity Task Force (CITF) grants VR-1 172711 and VS1-175545 (T.H.W. and A.C.G); CIHR FDN-143250 (T.H.W.), GA2-177716 (V.C., A.C.G., T.W.), GA1-177703 (A.C.G.) and the CIHR rapid response network to SARS-CoV-2 variants, CoVaRR-Net (to A.C.G.).

## Introduction

The COVID-19 pandemic caused by severe acute respiratory syndrome coronavirus 2 (SARS-CoV-2) remains a serious health crisis (1, 2). COVID-19 infections can vary from asymptomatic or mild through to severe disease, with lethal complications such as progressive pneumonia, acute respiratory distress syndrome and organ failure driven by hyperinflammation and a cytokine storm syndrome. Patients with immune-mediated inflammatory diseases (IMID), such as inflammatory bowel disease (IBD), psoriatic disease, rheumatoid arthritis (RA) and spondyloarthritis (SpA), are frequently treated with immunosuppressants and biologics and therefore may be at increased risk for COVID-19 (3, 4). Age and underlying comorbidities as well as the use of some immunosuppressants have been shown to be risk factors for developing COVID-19 among IMID patients (3, 5). Glucocorticoids and combination therapy of immunomodulators and biologics have been shown to increase the risk of severe outcomes of COVID-19 (4, 6).

Although many IMID patients mount adequate serological responses to vaccination after two doses of an mRNA vaccine, a proportion of IMID patients show reduced responses compared to healthy controls (7-14), as confirmed in recent meta-analyses (15, 16). In particular, patients receiving glucocorticoids, methotrexate, mycophenolate, anti-TNF and B-cell depleting therapy may have attenuated serological responses to COVID-19 vaccines (7, 11, 13, 15, 17, 18). A small study of 23 IMID patients showed that patients on anti-TNF therapy have greater waning of humoral immunity compared to healthy controls (13), as confirmed in a larger study still in preprint (19).

Data regarding the cellular immune responses to vaccination are still relatively scarce and conflicting. Several studies have shown unimpaired T cell responses to SARS-CoV-2 vaccines in immunocompromised patients compared to healthy individuals (13, 20-23), though a follow-up study showed that a proportion of IMID patients on immunosuppression had reduced T cell responses to a second dose of vaccine (24). In another study, methotrexate limited CD8^+^ T cell responses to vaccination in a cohort of IMID patients (25). To gain further insight into immunity to mRNA vaccines in IMID patients on different maintenance therapies, we investigated serological and T cell responses against SARS-CoV-2 before and after one or two doses of mRNA vaccine. The results show substantial variation in responses within different treatment groups. Notably, we observed decreased serological responses in anti-TNF treated patients, including decreased efficacy of neutralization of variants of concern, with no neutralizing capacity against the Omicron variant. T cell cytokine production, including IFN-γ, IL-2 and IL-4 increased from one to two doses of vaccine and correlated with humoral responses. Importantly, both antibody and T cell responses in the IMID treatment groups showed greater waning by 3 months post second dose of mRNA vaccine compared to healthy controls. These data highlight the need for third doses of SARS-CoV-2 mRNA vaccines and for continued monitoring of responses in these patients.

## Results

### Study population and design

Of 177 initially recruited subjects, 150 met the inclusion criteria for this study (see methods). PBMCs and plasma were collected for T cell and antibody responses at up to 4 time points, before and after vaccination with mRNA vaccines (Figure 1A). In our cohort, the median time between dose 1 and dose 2 of the mRNA vaccines was 60.5 days, IQR [45.5-72]. Baseline characteristics of the study subjects are shown in Table I. Of note, age and BMI, but not vaccine interval, were significantly different between groups and multivariate analysis of the data took these differences into account (Supplemental Tables S1-S3). The patients ultimately analyzed included 26 healthy controls, 9 IMID patients not on treatment, 44 IMID patients on anti-TNF, 16 on anti-TNF with MTX/AZA, 10 on anti-IL-23, 28 on anti-IL-12/23, 9 on anti-IL-17 and 8 on MTX/AZA.

**Figure 1.**
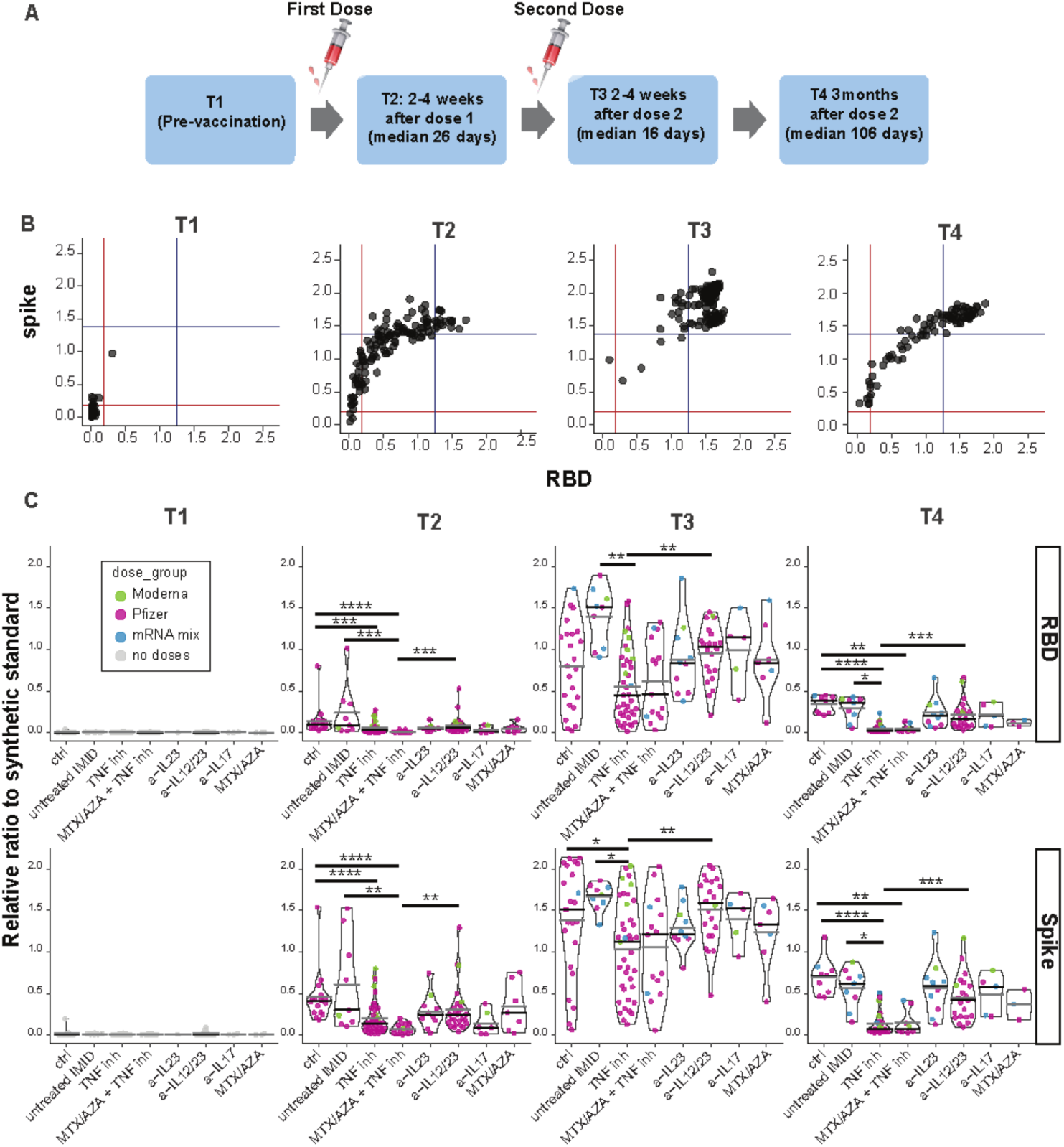
Antibody responses after 1 or 2 doses of mRNA vaccine. **(A)** Schematic diagram of the sampling schedule. **(B)** IgG response to two doses of vaccine across all participants. Anti-spike and anti-RBD IgG levels at indicated time points (defined in Figure 1A). The blue line is the median ratio in convalescent patients (340 samples collected 21 to 115 days post symptom onset), 1.38 and 1.25 for spike and RBD respectively. The red line is the seropositivity threshold: the median antibody level of those that pass both a 1% false positive rate (FPR) and show ≥3 standard deviations from the log means of the negative controls. **(C)** IgG responses before, and after the first and second doses of mRNA vaccine in IMID patients. Violin plots show the relative ratio of RBD and spike that were determined at the indicated time points in IMID patients under mono- and combination therapy (0.0039 µl sample used, see suppl. fig. 1 for the second dilution and suppl. table S1 for conversion to BAU/ml). T1, n=111; T2 n= 131, T3, n=131, T4, n= 88. The dot colors indicate the type of vaccine, Pfizer refers to BNT162b; Moderna to mRNA-1273; mRNA mix = first dose BNT162b, second dose mRNA-1273. MTX = methotrexate, AZA = azathioprine. Black and gray lines indicate median and mean ratio values for each violin, respectively. Plots are faceted based on the groups/treatments. Comparisons were made by Dunn’s multiple comparisons test based only on the BNT162b/BNT162b group. *\*p≤0.05, **p≤0.01, ***p≤0.001, ****p≤0.0001*

**Table I.** Characteristics of study participants at baseline and vaccination interval between two doses of an mRNA vaccine.

|  | control | untreated<br>IMID | Anti-TNF<br>agents | MTX/AZA +<br>anti-TNF<br>agents | anti-IL-23<br>agents | Anti-IL-12/23<br>agents | Anti-IL-17<br>agents | MTX/AZA |  |
| --- | --- | --- | --- | --- | --- | --- | --- | --- | --- |
|  | n=26 | n=9 | n=44 | n=16 | n=10 | n=28 | n=9 | n=8 | Sig. (p-value) |
| **IMID\*** | N/A |  |  |  |  |  |  |  |  |
| **IBD** |  | 9 | 30 | 10 | 0 | 27 | 0 | 4 |  |
| **Psoriasis** |  | 1 | 3 | 1 | 8 | 1 | 2 | 2 |  |
| **PsA** |  | 0 | 7 | 3 | 2 | 1 | 7 | 2 |  |
| **AS** |  | 0 | 8 | 3 | 0 | 0 | 1 | 0 |  |
| **RA** |  | 1 | 1 | 0 | 0 | 0 | 1 | 1 |  |
| **Age**<br>**median years**<br>**[IQR]** | 36 [26-46] | 33 [27-41] | 38 [30-51] | 53 [44-59] | 48 [45-61] | 34 [28-47] | 49 [46-61] | 42 [31-55] | <0.001^ |
| **Sex**<br>**male (%)** | 16 (62) | 5 (56) | 18 (41) | 8 (50) | 5 (50) | 13 (46) | 6 (67) | 4 (50) | 0.772~ |
| **BMI**<br>**median kg/m2**<br>**[IQR]** | 25 [23-28] | 26 [22-27] | 22 [24-26] | 26 [24-28] | 27 [24-35] | 22 [21-24] | 32 [26-34] | 26[25-33] | 0.001^ |
| **Vaccine interval**<br>**median days**<br>**[IQR]** | 74 [35-84] | 54 [31-64] | 60 [45-69] | 64 [50-72] | 74 [35-84] | 62 [49-69] | 65 [52-75] | 58 [21-97] | 0.372^ |

\*multiple IMIDs per patient possible ^Kruskal Wallis test. ~Fisher's exact test. \*\*1 patient in this study group was also on methotrexate. IMID; immune mediated inflammatory disease, TNF; tumor necrosis factor alpha, MTX, methotrexate; AZA, thiopurine, IL; interleukin, n; number, N/A; not applicable, IQR; interquartile range, BMI; body mass index, sig; significance, IBD; inflammatory bowel disease, PsA; psoriatic arthritis, RA: rheumatoid arthritis, AS; ankylosing spondylarthritis, kg/m2; kilogram per square mete

### Antibody responses are reduced in anti-TNF treated subjects and wane over time

Antibody responses were measured by automated ELISA. For the entire cohort, antibody responses increased from T1 to T2 to T3, and decreased by T4 (Figure 1B). Responses to NP were used to rule out exposure to SARS-CoV-2 (Supplemental Figure S1). After the first dose, 97.8% and 80% of participants seroconverted to spike and RBD IgG, respectively, and the relative ratios were greater than the medians of the convalescents in 44.2% and 14.2% of the participants (Figure 1C). Seroconversion increased to 100% for spike and 99.2% for RBD after the second dose and the anti-S and anti-RBD IgG levels were greater than the median levels of convalescent patients in 97% and 86.6% of participants, with a median relative ratio of 1.91 for spike and 1.55 for RBD. Analysis of antibody responses by vaccine type showed that two doses of the mRNA-1273 vaccine elicits a stronger humoral response than BNT162b, with mixed mRNA vaccines inducing significantly higher levels of anti-spike/RBD IgG than two doses of BNT162b (Supplemental Fig. S2A). Although all data were included in the figures, as the majority of the cohort was vaccinated twice with BNT162b2, univariate statistical analysis between treatment groups was performed only on samples from the BNT162b/BNT162b participants. Among the BNT162b/BNT162 cohort, males had a slightly lower response to RBD than females, whereas antibody response differences by age were not significant (Supplemental figure S2B, C). Participants undergoing anti-TNF, and anti-TNF+MTX/AZA therapies had significantly lower levels of antibodies than those in the healthy control, IMID-untreated, and anti-IL-12/23 groups after the first dose of vaccine (Figure 1C and Supplemental Fig. S1). Comparison between the groups after the second dose (T3) indicates that for the BNT162b/BNT162b group, participants taking anti-TNF had significantly lower levels of anti-spike IgG than those in the healthy control, IMID-untreated, and anti-IL-12/23 groups (Figure 1C). Multivariate analysis of treatment groups controlling for age and sex confirmed the deficits in anti-RBD and anti-spike in the anti-TNF group after the second dose, whether the entire cohort or only the BNT162b/BNT162b participants were evaluated (Suppl. Table S1).

Spike and RBD antibody levels decreased by T4 (median 106 days post-dose 2), with a more rapid decline in anti-RBD levels (Figure 1C). Only 66.3% and 49.4% of the participants show relative ratios greater than the medians of the convalescents for spike and RBD, respectively. When data were ranked by study group, we observed that the anti-TNF, and anti-TNF+MTX/AZA therapy groups were associated with a statistically significant drop in anti-spike and anti-RBD IgG levels compared to the healthy control, IMID-untreated, and anti-IL-12/23 groups (Figure 1C, Supplemental Table S1). Multivariate analysis of treatment groups controlling for age and sex confirmed the deficits in anti-RBD and anti-spike in the anti-TNF group at T4, whether the entire cohort or only the BNT162b/BNT162b participants were evaluated (Supplemental Table S1).

### IMID patients undergoing anti-TNF therapy show significantly lower neutralization responses than other groups

We performed at T3 and T4 (2-4 weeks and 3 months after the second vaccine dose) spike-pseudotyped lentiviral neutralization assays using the wild-type strain and B.1.351 (Beta), P.1 (Gamma), B.1.617.2 (Delta), and/or B. 1.1.529 (Omicron) variants of concern (VOCs). At T3, samples neutralized the wildtype more efficiently than the VOCs tested, but participants on anti-TNF and anti-TNF+MTX/AZA showed significantly lower neutralization response to all variants (Figure 2A & Supplemental Figure S3). Consistent with the waning antibody levels at T4, median neutralization was reduced for the wild-type and Delta lentiviruses (Supplemental Figure S3), and again, the participants on anti-TNF and anti-TNF+MTX/AZA showed significantly lower neutralization response (Figure 2B). Consistent with recent reports,(26-30) Omicron was about an order of magnitude more difficult to neutralize than wild-type and Delta spike-pseudotyped lentiviral particles, and sera from anti-TNF and anti-TNF+MTX/AZA were unable to produce detectable neutralization in our assay (Figure 2B).

**Figure 2.**
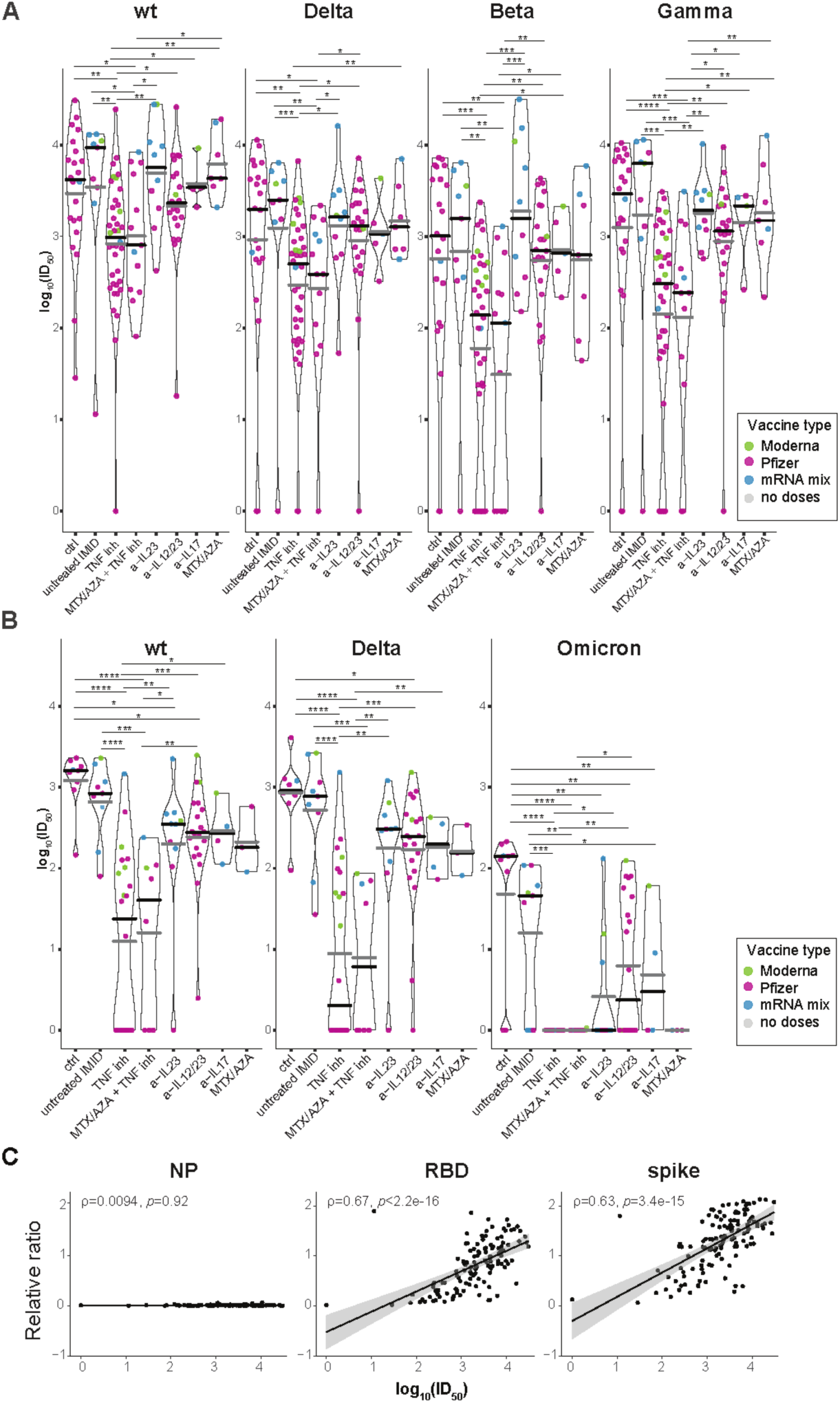
Variant neutralization after two doses of vaccine. **(A & B)** Violin plots of log10 ID50 (serum dilution that inhibits 50% of the infectivity) values of samples at A, time point 3 (2-4 weeks post dose 2), n=129 and B, time point 4 (3 months post dose 2), n=86 (see Figure 1A). Lentiviral particles used: wildtype, B.1.617.2 (Delta), B.1.351 (Beta), P.1 (Gamma), and B. 1.1.529 (Omicron). The distribution is stratified by study groups/treatments. The dots colors indicate the type of vaccine. Black lines indicate the median and gray lines the mean ratio value for each violin. Comparisons were made by Dunn’s multiple comparisons test for the entire cohort. *\*p≤0.05, **p≤0.01, ***p≤0.001, ****p≤0.0001*. **(C)** Correlation between the wild-type neutralization responses and IgG levels using the log10 ID50 values of neutralization and the relative ratio of the spike and RBD IgG levels at T3 (supplemental Figure S3 for correlations with the VOCs).

Overall, these data demonstrate weaker neutralization responses to mRNA vaccines at the time points tested for the participants on anti-TNF agents. Consistent with the fact that the same trends were detected in the ELISA data, moderate Spearman’s correlations (ρ = 0.59-0.67) were detected between anti-spike/RBD IgG levels and lentiviral neutralization of the wild-type and VOCs tested (Figure 2C and Supplemental Figure S3C), though a subset of participants had high levels of anti-spike/RBD IgG failed to neutralize viral entry.

### IMID patients show increased T responses to successive vaccine doses, with greater waning after dose 2

To assess memory T cell responses to SARS-CoV-2, PBMCs were stimulated with spike or NP peptide pools for 48hrs. A quantitative multiplex bead-based immunoassay was used to measure the levels of 9 secreted cytokines and cytotoxic molecules in the supernatants in response to spike peptide stimulation and results are reported after subtracting the values from negative control wells. The response to NP was used as an additional control to detect memory responses to previous virus exposure. NP-specific responses pre-vaccination were minimal, consistent with study subjects being SARS-CoV-2 naïve and suggesting minimal impact of cross-reactive T cells from previous coronavirus infections (Supplemental Figure S4). The cytokines IFN-γ, IL-2, IL-17A and IL-4 were increased over baseline (T1) after one or two doses of mRNA vaccine in all patient groups (T2 and T3), with the response predominantly of the Th1 phenotype as characterized by high levels of IFN-γ and IL-2 (Figure 3, Supplemental figure S5). Molecules associated with cytotoxicity such as granzyme (Gzm) A, GzmB, perforin and sFasL were also increased over baseline following one dose of vaccine and did not consistently increase with the second dose (Figure 4, Supplemental Figure S5). TNF was not detected over baseline (data not shown). Most study groups showed a wide range of responses to spike peptide pools after first or second vaccine doses (Figures 3, 4, Supplemental Figure S5) When multivariate analysis was performed on the BNT162b/BNT162b group only, after controlling for age/sex/BMI, we observed deficits in IL-2 or IFN-γ production in IMID untreated, anti-TNF, anti-IL-17, MTX, and anti-IL-12/23 treatment groups relative to healthy controls after dose one, which largely recovered by dose two (Supplemental Table S1). However, by T4, T cell responses were lower in most treatment groups of IMID patients as well as IMID patients relative to controls (Supplemental Table S1). When results from all subjects were pooled, there was an increase in response from first to second dose for all 8 readouts (Figure 5A, B). By 3 months after dose 2 (T4), we saw an overall decrease in IL-2, IL-4, IL-17, sFASL and Granzyme A (Figure 5A, B). We also noted higher IL-4 responses following vaccination with mRNA-1273 compared to BNT162b or mixed doses (Supplemental Figure S6A). Although T cell responses overall were similar based on age or sex (suppl. fig. S6B, C), multivariate analysis revealed lower IL-4 responses in the over 60 group (Supplemental Table S2).

**Figure 3.**
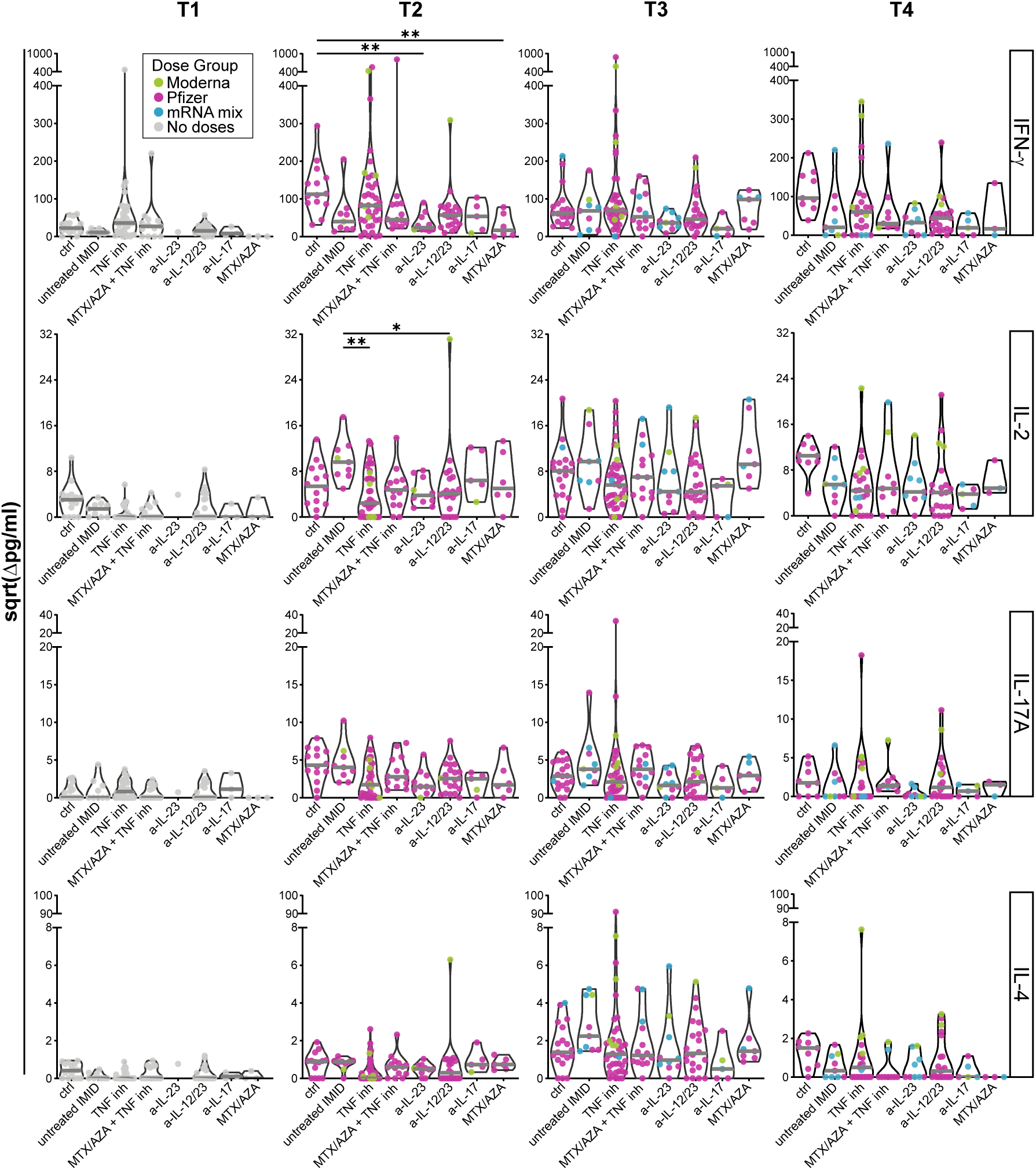
Cytokine responses in each group prior to vaccination and after first and second doses of mRNA vaccine. Cytokine release in cell culture supernatants was analyzed by multiplex bead array following 48h stimulation with SARS-CoV-2 spike peptide pools. Violin plots show IFN-γ, IL-2, IL-17A and IL-4 release at T1 (pre-vaccination); n=102, T2; n=117, T3; n=126, and T4; n= 88, with timepoints defined in Figure 1A. Colored dots represent the type of vaccine (defined in figure 1B). The gray line indicates the median. Values are reported in pg/ml after subtracting background signal from wells containing PBMCs cultured with DMSO alone, as indicated by “Δ”. Ctrl = Healthy controls, inh = inhibitor, MTX = methotrexate, AZA = thiopurines. Comparisons between groups in entire cohort were made by Dunn’s multiple comparisons test after excluding outliers and subjects with an NP IgG response. *\*p≤0.05, **p≤0.01*.

**Figure 4.**
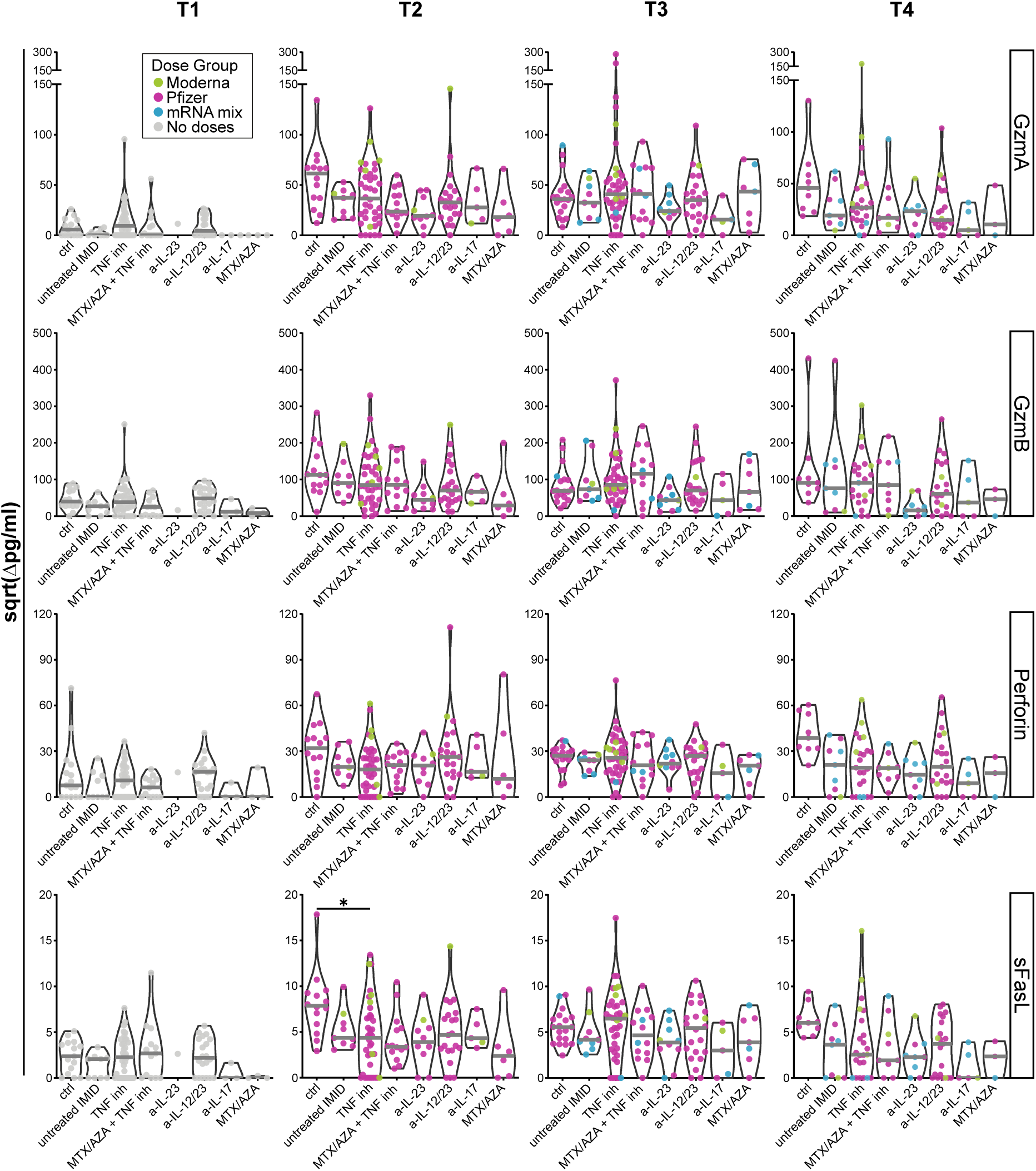
Cytotoxic responses in each group before or after first and second doses of mRNA vaccine. The release of cytotoxic molecules in cell culture supernatants was analyzed by multiplex bead array following 48h stimulation with SARS-CoV-2 spike peptide pools. Violin plots show release of Granzyme (Gzm) A, B, perforin or sFASL release at T1, n=102; T2, n=117; T3, n=126, and T4, n= 88 (with T1-T4 defined in figure 1A). The dot colors indicate the type of vaccine (see figure 1B). The gray line indicates the median. Values are reported in pg/ml after subtracting background signal from wells containing PBMCs cultured with DMSO alone, as indicated by “Δ”. Ctrl = Healthy controls, inh = inhibitor, MTX = methotrexate, AZA = thiopurines. Comparisons on entire cohort were made by Dunn’s multiple comparisons test after excluding outliers and subjects with an NP IgG response. *\*p≤0.05*.

**Figure 5.**
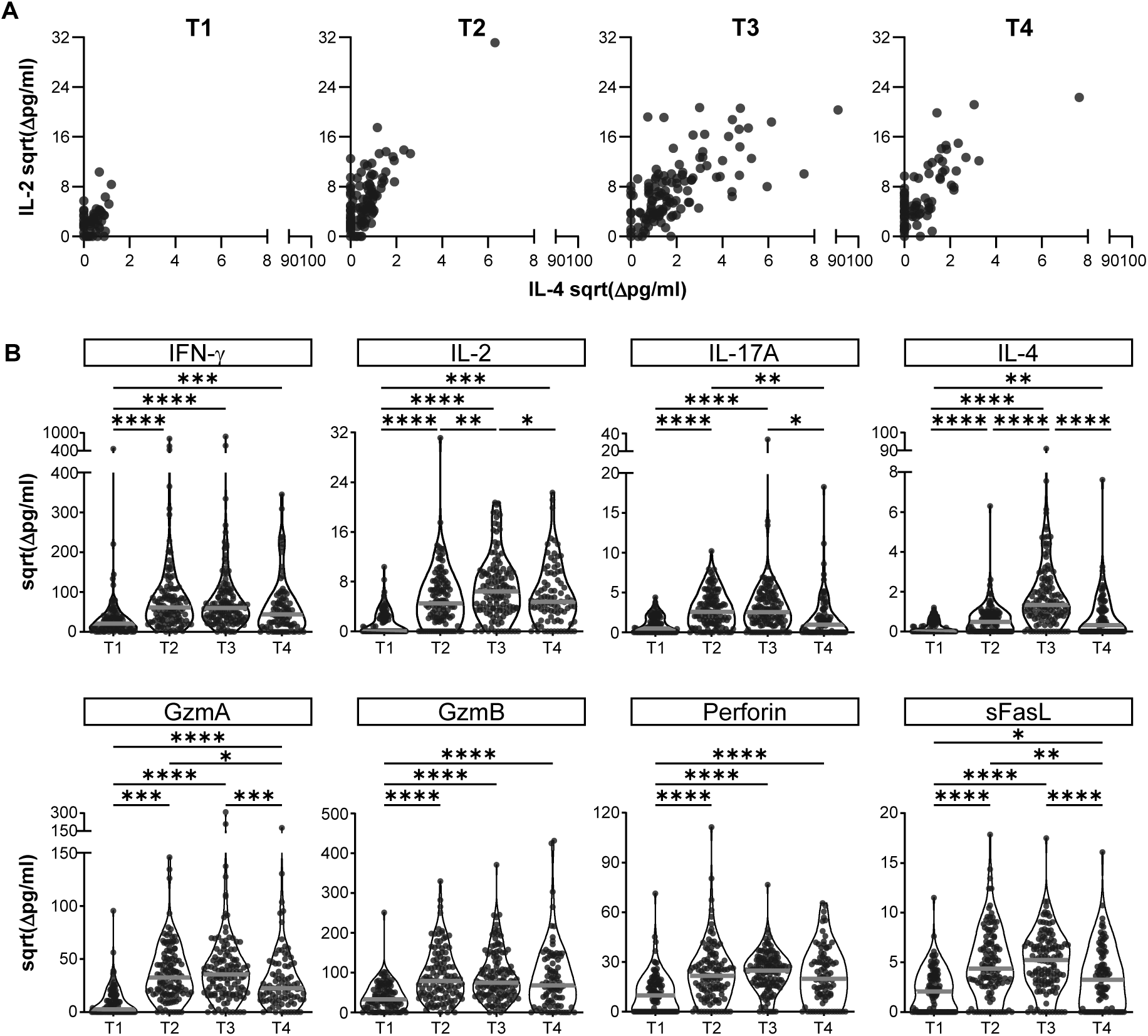
Cytokine and cytotoxic responses in all patient groups in response to Spike peptide pools over time. The release of cytokines in cell culture supernatants were analyzed by multiplex bead array following 48h stimulation with SARSCoV-2 S peptide pools. **(A)** IL-2 and IL-4 responses across all participants at timepoints T1 -T4 as defined in figure 1A. **(B)** Violin plots show release of cytokines and cytotoxic molecules in all study subjects pooled. The median is indicated by the grey line. Pairwise comparisons were made by mixed-effects ANOVA after excluding outliers and subjects with an NP IgG response. *\*p≤0.05, **p≤0.01, ***p≤0.001, ****p≤0.0001*.

Levels of secreted IL-2 were positively correlated with plasma IgG against RBD (r=0.50) and whole spike trimer (r=0.51). Similarly, there was a positive correlation between IL-4 and plasma IgG against RBD (r=0.58) and whole spike trimer (r=0.59), and between IFN-γ and RBD IgG (r=0.36) and whole spike trimer IgG (r=0.36) (all p values <0.0001) (Figure 6).

**Figure 6.**
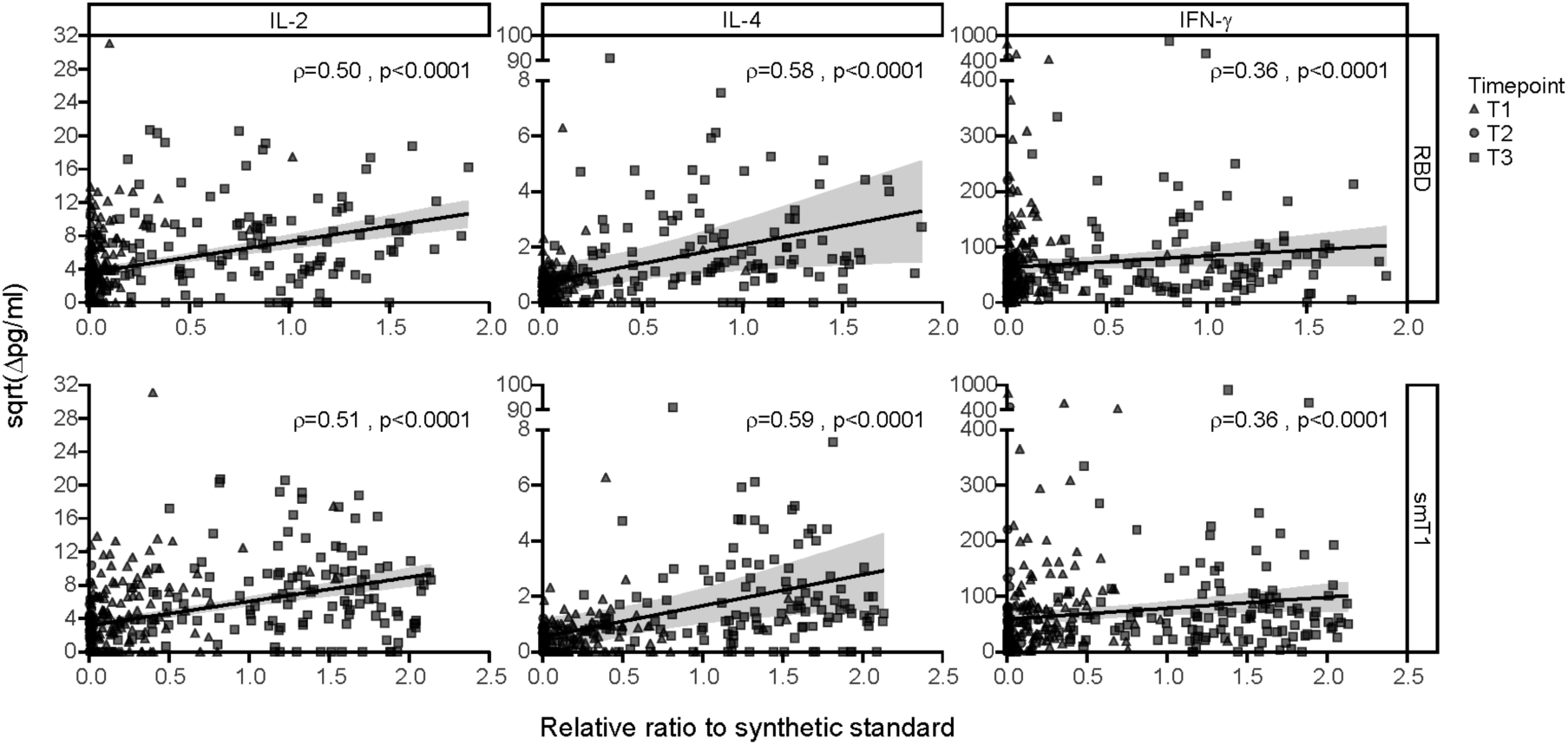
Correlation between IgG levels and T cell cytokine responses at all sampled time points. The solid black line is the linear regression, and the gray shading indicates the 95% confidence interval. p-values and Spearman’s rho coefficients are indicated in each graph.

## Discussion

Here we studied a cohort of patients with inflammatory bowel disease, psoriatic disease, ankylosing spondylitis or rheumatoid arthritis, treated with biologics (anti-TNF, anti-IL-12/23, anti-IL-23, anti-IL-17) or antimetabolites to assess their response to COVID-19 mRNA vaccines. Although this group is not considered to be significantly immunosuppressed, there has been concern as to how their treatments could impact the response to the vaccines. Although there was considerable variability within groups, 100% of participants seroconverted for spike after 2 doses of vaccine. There was also a clear indication of higher responses to mRNA-1273 vaccine compared to BNT162b vaccine with respect to antibody levels and neutralization titers, as well as T cell IL-4 production. Of concern, antibody levels and neutralization activity were lower in the anti-TNF treated study subjects even after two doses of vaccine and showed accelerated waning by 3 months post dose 2, with neutralization of the Omicron S-pseudotyped lentivirus undetectable in this group at that time point. Our data are consistent with recent data suggesting reduced vaccine efficacy against Omicron infection in immunocompromised patients (14). The observed waning of antibody responses to mRNA vaccines in anti-TNF patients are in agreement with a recent small study from Geissen et al. (13) who showed decreased responses and waning immunity with anti-TNF agents in 23 IMID patients at 6 months post dose 2, and also confirmed with a larger cohort in a recent pre-print (19). We also observed that IMID patients overall showed more substantial waning of both antibody and T cell responses compared to healthy controls. In our study, the vaccine dose interval was a median of 60.5 days rather than the standard 21 or 28 days used in the other studies, which could impact the results. Some limitations of our study are the small numbers of study subjects in some of the groups and grouping together of drugs by class.

T cell responses, including IL-4, IL-2 and IFN-γ production, showed a significant correlation with RBD and spike - specific antibody responses. There was substantial induction of T cell cytokines and release of cytotoxic molecules following spike peptide pool stimulation of PBMCs collected following one dose of vaccine and this increased further for all readouts after two doses. Multivariate analysis of the data showed that several groups had decreased IFN-γ after dose 1 of vaccine, but these deficits were largely corrected following the second vaccine dose. When data were pooled for all subjects, it was apparent that cytokine responses and IL-4 and IL-17 in particular, were dependent on 2 doses of vaccine. IL-4 is an important mediator of B cell proliferation, which in turn impacts antibody levels and B cell memory (31). This lower IL-4 response after 1 dose as compared to 2 doses of vaccine highlights the need for second doses to maximize B cell responses. Of note, a recent report showed that atopic dermatitis as well as asthma patients treated with either IL-4 or IL-5 receptor antagonists had reduced antibody responses following two doses of mRNA vaccines compared to healthy controls, consistent with the importance of T cell IL-4 in the antibody response to SARS-CoV-2 mRNA vaccines (32).

Taken together, our study shows generally robust T cell responses in most patient groups treated with immunosuppressants or biologics after 2 doses of mRNA vaccine, improving with a second dose but with significantly more attenuation in IMID patients than healthy controls by 3 months after the second dose. We observed substantial deficits in antibody responses even after two doses of vaccine in the anti-TNF treated patients, with more substantial waning immunity by three months after dose two and a complete inability to neutralize the Omicron variant. These findings highlight the need for a third vaccine dose, particularly in patients undergoing treatment with anti-TNF agents. As there is limited information available about the duration of immune memory induced by mRNA vaccines, it will also be important to follow these responses for longer time periods and to evaluate the impact of additional vaccine doses in this cohort as well as the possible contribution of natural infection to persistence of immune response.

## Methods

### Study design and participants

Patient recruitment: In this observational multicenter cohort study, we investigated the IMmune resPonse After COVID-19 vaccination during maintenance Therapy in immune-mediated inflammatory diseases (IMPACT). IMID patients being treated at Mount Sinai Hospital, University Health Network/Toronto Western Hospital or Women’s College Hospital in Toronto, Canada who were receiving BNT162b (Pfizer-BioNTech) and/or mRNA-1273 (Moderna) SARS-CoV-2 vaccines were recruited between January 8 and October 4, 2021. It should be noted that in Canada the vaccine schedule between dose 1 and 2 was increased from the standard 21 or 28 days to allow faster roll out of dose 1 and as a result, in our cohort there was a median of 60.5 days, IQR [45.5-72] between the 2 doses.

Inclusion criteria for this study were adult IMID patients being treated with anti-TNF therapies (infliximab, adalimumab, golimumab, etanercept or certolizumab pegol), anti-IL-17 therapy (ixekizumab, secukinumab), methotrexate (MTX) or azathioprine (AZA) monotherapy, combination therapy of MTX/AZA plus anti-TNF therapies, anti-IL-12/23 (ustekinumab) therapy, anti-IL-23 therapy (guselkumab, risankizumab) or no immunosuppressants. A group of healthy volunteers, without an IMID and without immunosuppression were also recruited as a control cohort. Excluded were individuals younger than 18 years, those who had a past SARS-CoV-2 infection, patients on vedolizumab or oral steroids and those receiving COVID-19 vaccines other than mRNA.

Sample and data collection: Patient information and medical history were collected at each visit. Participation was terminated when all the blood samples were collected or when a patient opted out. Clinical data included basic demographics (age, sex, weight, height), relevant past medical and surgical history, and medication use at inclusion. Questions about prior COVID-19 diagnosis or exposure, vaccination history and side effects, changes in medical history or medication and disease activity were collected at each study visit. Blood samples were drawn from the participants at up to 4 time points: T1 = pre-vaccination, T2 = median 26 days after dose 1, T3 = median 16 days after dose 2 and T4 = median 106 days after dose 2. Peripheral blood samples were collected in BD Vacutainer^®^ sodium heparin tubes for plasma antibody assessment and peripheral blood mononuclear cell (PBMC) separation. All samples were labelled with unique patient identifiers. Researchers were blinded to the identity and clinical details of the subjects. Plasma samples were stored at -80°C. PBMCs were isolated by density centrifugation using Ficoll-Paque PLUS (GE Healthcare). PBMCs were cryopreserved in 10% DMSO in FBS (Wisent Bioproducts) and stored in liquid nitrogen at a minimum of 2×10^6^ mononuclear cells per vial.

### Automated ELISAs

Frozen plasma was thawed and treated with 1% final Triton X-100 for one hour. Samples were analyzed by automated ELISA for IgGs to the spike trimer (spike), the spike receptor binding domain (RBD), and the nucleocapsid (NP; all antigens and secondary antibodies are produced in mammalian cells and were provided by Dr. Yves Durocher at the National Research Council of Canada, NRC) as previously reported (33). Luminescence values for each sample/assay were normalized to synthetic standards profiled in a 4-fold dilution series on each plate (Human anti-nucleocapsid IgG, #A02039, clone HC2003, GenScript, Piscataway, NJ, USA and humanized anti RBD/spike IgG: VHH72hFc1X7; NRC). The synthetic references, as well as a pool of positive samples from convalescent patients with high IgG level to all three antigens and negative controls (pre-COVID era samples, blank and IgG, 1 µg/ml; #I4506, Millipore-Sigma, Oakville, ON, Canada) were also added to each plate in a 4-fold dilutions series to enable quality controls across the plates and batches of samples. For each assay, log10 raw values and relative ratio of samples were compared to prior runs to confirm that the sample density distribution is within range; automated scripts, blinded to sample description and meta-data were used to extract relative ratios to the synthetic references. The assay was calibrated to the World Health Organization (WHO) reference (National Institute for Biological Standards and Control, NIBSC, Code 20/136); a table of conversion of relative ratios for each assay to Binding International Units/ml (BAU/ml) is provided (suppl. table S1). Seropositivity was defined based on both receiver operating characteristic (ROC) analysis of negative (pre-COVID era) and positive (PCR confirmed COVID-19 cases) samples (<1% false positive rate threshold) and on deviation from the log means of the negative controls (≥ 3 standard deviations). In some of the figures, the median convalescent values for serum samples from 340 PCR confirmed COVID-19 cases 21–115 days after symptom onset(33) is displayed as a reference point. Since the assays saturate in healthy controls after two doses of vaccine, all samples were processed both at the dilution used for determination of seroconversion, and a 1/16 further dilution for evaluation of the quantitative differences in antibody responses.

### Spike-pseudotyped lentivirus neutralization assays

The lentivirus neutralization assay and the generation of spike pseudotyped lentivirus particles were performed as described previously.(34) Briefly, the lentivirus particles were generated by co-transfection in HEK293TN cells (System Biosciences, Palo Alto, CA, USA, LV900A-1) of the Wuhan Hu-1 sequence with a D614G mutation (wild-type SARS-CoV-2), or the variants B.1.617.2 (Delta), B.1.351 (Beta), P.1 (Gamma), and B. 1.1.529 (Omicron constructs with packaging (psPAX2, Addgene, Watertown, MA, USA, #12260) and reporter (luciferase expressing pHAGE-CMV-Luc2-IRES-ZsGreen-W, provided by Drs. Jesse Bloom and Katharine Crawford) constructs. Heat-inactivated (30 min at 56°C) plasma was serially diluted and incubated with the lentiviral particles (1h, 37°C) prior to addition to cells (HEK293T-ACE2/TMPRSS2) for 48h; luminescence signals were detected with the Bright-Glo Luciferase assay system (Promega, E2620) on an EnVision multimode plate reader (Perkin Elmer. GraphPad Prism 9 was used to calculate 50% neutralization titer (ID50) using non-linear regression. The WHO International Standard (20/136) was evaluated in this assay, and a mean ID50 value of 5744 corresponded with 1000 IU/ml.

### T cell cytokine secretion assay

Cellular immune responses to COVID-19 vaccination were determined by measuring the release of cytokines and cytotoxic molecules in cell culture supernatants following stimulation with peptide arrays using the LEGENDplex multiplex bead assay as previously described (35, 36). Briefly, 1×10^6^ PBMCs were seeded per well in 96-well round bottom plates with 1 µg/ml each of SARS-CoV-2 spike or Nucleoprotein (NP) peptide pools (JPT peptide technologies, GMBH, Berlin, Germany). PBMCs were cultured with anti-CD28 (clone 9.3, Bio X Cell) and anti-CD3 (clone OKT3, Bio X Cell) as a positive control, or with equimolar DMSO as a negative control. Samples with no response to positive control were not included in the analysis. After 48h incubation at 37°C, cell culture supernatants were collected and stored at -80°C. Release of cytokines and cytotoxic molecules (IL-2, 4, 17, IFN-γ, TNF, Granzyme A,B, Perforin, sFASL) in the supernatants were analyzed using LEGENDplex multiplex cytokine bead assay (BioLegend) as per manufacturer’s instructions. Samples were acquired on the BD LSR Fortessa flow cytometer using BD FACSDiva software. Data are reported as square root (sqrt) transformed values in pg/ml after subtracting background signal from wells containing PBMCs cultured with DMSO containing media alone, as indicated by “Δ”.

### Statistical analysis

T cell cytokine secretion data were analyzed using the LEGENDplex(tm) Data Analysis Software Suite, pandas data analysis library for Python and GraphPad Prism v9.3.1 (37). Antibody data were analyzed with R (version 4.1.1) using package ggplot2 and custom R scripts. GraphPad Prism v 9.2.0 was used to analyze the neutralization and antibody data. Longitudinal multivariate analysis on antibody data and T cell cytokine secretion was performed using linear mixed models. Models controlled for baseline (timepoint 1) T cell/antibody data and included an interaction term between time point and the variable of interest. All multivariate analyses were performed using R (version 4.1.1) and SAS 9.4.

### Study approval

This study was approved by the ethics boards of the University of Toronto (REB protocol #27673), Mount Sinai Hospital/Sinai Health System (MSH REB #21-0022-E), University Health Network-Toronto Western Hospital division (REB # 21-5096) and Women’s College Hospital (REB approval 2021-0023-E). Written informed consent was obtained from all participants prior to participation.

## Data Availability

All data in the present study are available upon reasonable request to the authors

## Author contributions

RMD and JCL shared first authorship, order was decided by mutual agreement

MSS and THW conceived the study and obtained funding.

ACG and THW supervised the laboratory assays, analyzed data, acquired funding, and wrote the manuscript.

RDI, NH, VP, VC and MSS contributed to study design, acquisition of funding, supervised clinical coordinators, contributed to data interpretation and manuscript editing.

RMD performed neutralization assays, analyzed all antibody data, conducted statistical analysis, prepared figures, and wrote the manuscript.

JCL designed and performed T cell assays, analyzed all T cell data, conducted statistical analysis, prepared figures, and wrote the manuscript.

RLG contributed to study conception and design, data analysis, literature review, and wrote manuscript

GYCC provided overall project management including patient recruitment and validation of data records.

MS conducted statistical analysis.

NVB, MG, IL, RL, MWC assisted with PBMC preparation and/or T cell experiments.

BR was responsible for sample intake and ELISA assays.

RS and QH generated and titrated the lentiviral stocks and optimized the neutralization assays for VOCs.

WRH generated all spike pseudotyped lentiviral vectors

KTA and JK wrote scripts for data analysis of the antibody data and generated figures.NF contributed to patient recruitment, literature review and manuscript preparation.

DC contributed to patient recruitment.

JS, DP, LA, SR, RM, KR, DG contributed to study coordination, and patient recruitment.

## Acknowledgements

We thank Juan and Stefania Speck for their generous donation to the University of Toronto for this study. We thank Drs. Jesse Bloom and Katharine Crawford for the initial spike lentiviral construct. All antigens and protein reagents for the automated ELISAs were a kind gift from Dr. Yves Durocher at the National Research Council of Canada (NRC) generated within the NRC’s Pandemic Response Challenge Program. We thank all members of the serology team at the Network Biology Collaborative Centre for help with ELISA assay development and automated ELISA processing, and in particular Drs. Karen Colwill and Adrian Pasculescu for providing the BAU/ml conversion table. We thank Birinder Ghumman for technical assistance and Nathalie Simard and Janine Charron for flow cytometry support. We thank the Royal Bank of Canada and the Krembil Foundation to the Sinai Health System Foundation for generous dontations to fund initial assay development in the Gingras lab. The calibration of the automated assays was supported notably through the COVID-19 Immunity Task Force (CITF). The robotics equipment used is housed in the Network Biology Collaborative Centre at the LTRI, a facility supported by the Canada Foundation for Innovation, the Ontario Government, and Genome Canada and Ontario Genomics (OGI-139). Anne-Claude Gingras is the Canada Research Chair in Functional Proteomics and the lead of the functional genomics and structure-function pillar of CoVaRR-Net. Tania Watts holds the Canada Research Chair in anti-viral immunity at the University of Toronto. Vinod Chandran is supported by a supported by a Pfizer Chair Research Award, Rheumatology, University of Toronto, Canada. Jaclyn C. Law was supported by an Ontario Graduate Scholarship. Kento T. Abe and Melanie Girard were supported by CIHR CGS-D studentships and Julia Kitaygorodsky is supported by a National Science and Engineering Research of Canada studentship.

