## Supplementary Figures for "Accelerated waning of immunity to SARS-CoV-2 mRNA vaccines in patients with immune mediated inflammatory diseases"

**Supplementary materials**

Supplemental Table S1-S3 Multivariable linear mixed model for treatment group, age and BMI are attached separately as excel files.

**Table S4  BAU/mL conversions**

**Nucleocapsid - Plasma**

| **Dilution (d)** | | **160** | | **2,560** | | |
|---|---|---|---|---|---|---|
| Volume equivalent (µl) | | 0.0625 | | 0.00391 | | |
| Relative Ratio (RR) | $log_2$(RR) | $log_2$(BAU/ml) | BAU/ml | $log_2$(BAU/ml) | BAU/ml | |
| 2 | 1.0 | 8.38 | 333.98 | 12.38 | 5,343.76 | Upper limit of linear range |
| 1 | 0.0 | 6.98 | 126.34 | 10.98 | 2,021.37 | |
| 0.5 | -1.0 | 5.58 | 47.79 | 9.58 | 764.62 | |
| 0.396 | -1.3 | 5.16 | 34.46 | 9.16 | 571.19 | Positivity threshold 160-fold dilution (0.0625 µl/well) |
| 0.25 | -2.0 | 4.18 | 18.08 | 8.17 | 289.23 | |
| 0.125 | -3.0 | 2.77 | 6.84 | 6.77 | 109.41 | |
| 0.0625 | -4.0 | 1.37 | 2.59 | 5.37 | 41.38 | Lower limit of linear range |
| **$log_2$(BAU/mL @ sample dilution d) = ($log_2$(RR) - 0.243)/ 0.713 + $log_2$(d)**<br>**With standard volume equivalents of .0625 µl and .00391 µl, range is 3 to 5344 BAU/ml, with positivity threshold of 34 BAU/ml** | | | | | | |

**RBD - Plasma**

| **Dilution (d)** | | **160** | | **2,560** | | |
|---|---|---|---|---|---|---|
| Volume equivalent (µl) | | 0.0625 | | 0.00391 | | |
| Relative Ratio (RR) | $log_2$(RR) | $log_2$(BAU/ml) | BAU/ml | $log_2$(BAU/ml) | BAU/ml | |
| 1 | 0.0 | 8.12 | 278.37 | 12.12 | 4,453.99 | Upper limit of linear range |
| 0.5 | -1.0 | 6.82 | 112.63 | 10.82 | 1,802.02 | |
| 0.25 | -2.0 | 5.51 | 45.57 | 9.51 | 729.07 | |

| | | | | | | |
|---|---|---|---|---|---|---|
| 0.186 | -2.4 | 4.95 | 30.97 | 8.95 | 495.58 | Positivity threshold 160-fold dilution (0.0625 µl/well) |
| 0.125 | -3.0 | 4.20 | 18.44 | 8.20 | 294.97 | |
| 0.0625 | -4.0 | 2.90 | 7.46 | 6.90 | 119.34 | |
| 0.03125 | -5.0 | 1.59 | 3.02 | 5.59 | 48.28 | Lower limit of linear range |
| **$\log_2$(BAU/ml @ sample dilution d) = ($\log_2$(RR) + 0.612)/ 0.766 + $\log_2$(d).** **With standard volume equivalents of .0625 µl and .00391 µl, range is 2 to 4,454 BAU/ml, with positivity threshold of 31 BAU/ml** | | | | | | |

### Spike - Plasma

| **Dilution (fold)** | | **160** | | **2,560** | | |
|---|---|---|---|---|---|---|
| Volume equivalent (µl added per well) | | 0.0625 | | 0.00391 | | |
| Relative Ratio (RR) | $\log_2$(RR) | $\log_2$(BAU/ml) | BAU/ml | $\log_2$(BAU/ml) | BAU/ml | |
| 1 | 0.0 | 6.55 | 93.80 | 10.55 | 1,500.80 | Upper limit of linear range |
| 0.5 | -1.0 | 5.28 | 38.75 | 9.28 | 619.95 | |
| 0.25 | -2.0 | 4.00 | 16.01 | 8.00 | 256.09 | |
| 0.19 | -2.4 | 3.50 | 11.28 | 7.50 | 180.45 | Positivity threshold at 160-fold dilution (0.0625 µl/well) |
| 0.125 | -3.0 | 2.72 | 6.61 | 6.72 | 105.78 | |
| 0.0625 | -4.0 | 1.45 | 2.73 | 5.45 | 43.70 | |
| 0.03125 | -5.0 | 0.17 | 1.13 | 4.17 | 18.05 | Lower limit of linear range |
| **$\log_2$(BAU/ml @ dilution d) = ($\log_2$(RR) - 0.604)/0.784 + $\log_2$(d)** **With standard volume equivalents of .0625 µl and .00391 µl, range is 1 to 1,501 BAU/ml, with positivity threshold of 11 BAU/ml** | | | | | | |

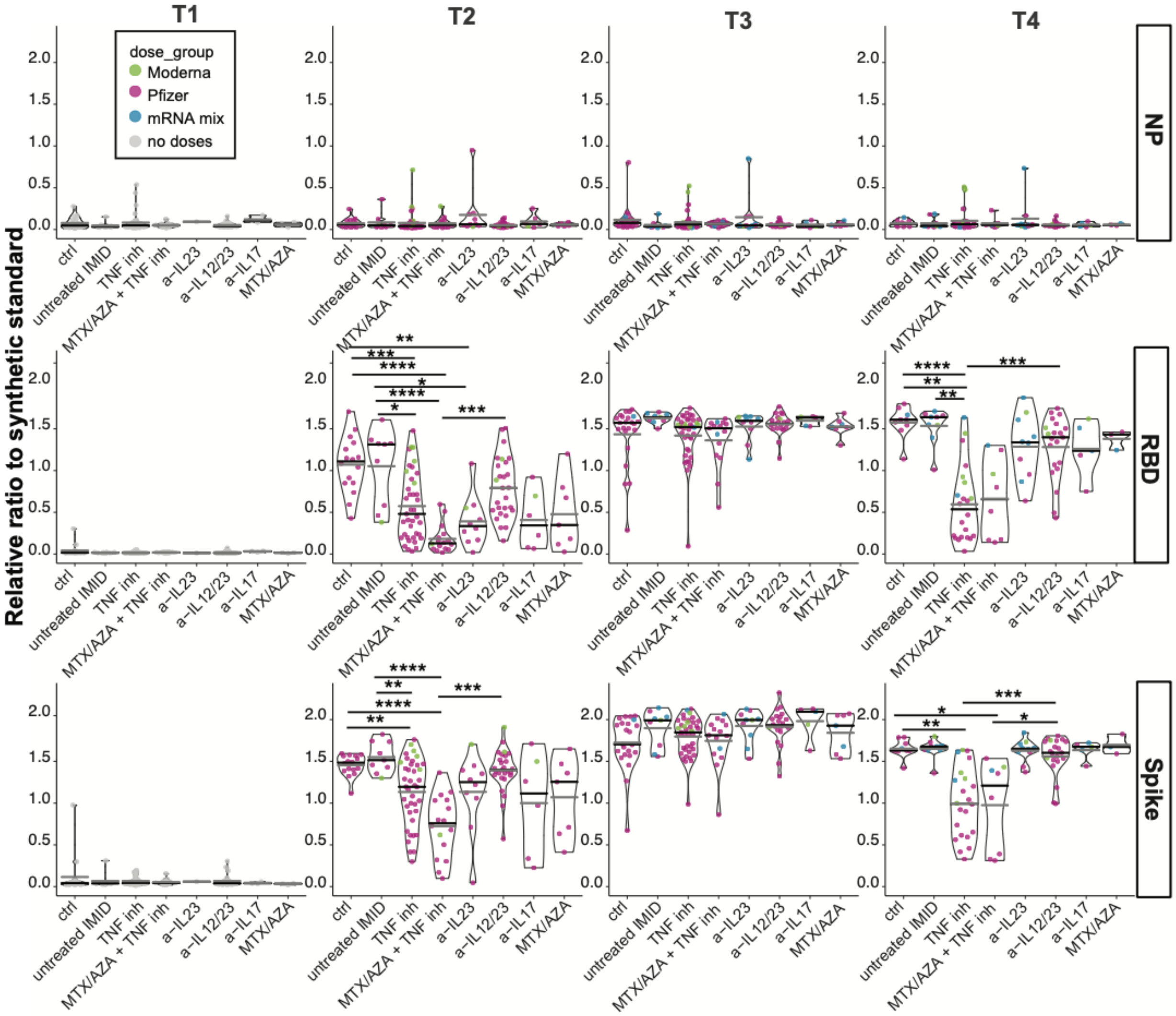


**Figure S1.** Antibody responses. Relative ratio of RBD, spike and nucleocapsid (NP) across time points in IMID patients on mono and combination therapy (0.0625 µl sample used). Dot colors represent the type of vaccine, Pfizer (BNT162b2), Moderna (mRNA-1273) and mRNA mix (first dose BNT162b, second dose mRNA-1273), respectively. Black and gray lines indicate median and mean ratio values, respectively. Statistical assessment by Dunn's multiple comparisons test was restricted to the BNT162b/BNT162b cohort. **p≤0.05, **p≤0.01, ***p≤0.001, ****p≤0.0001*

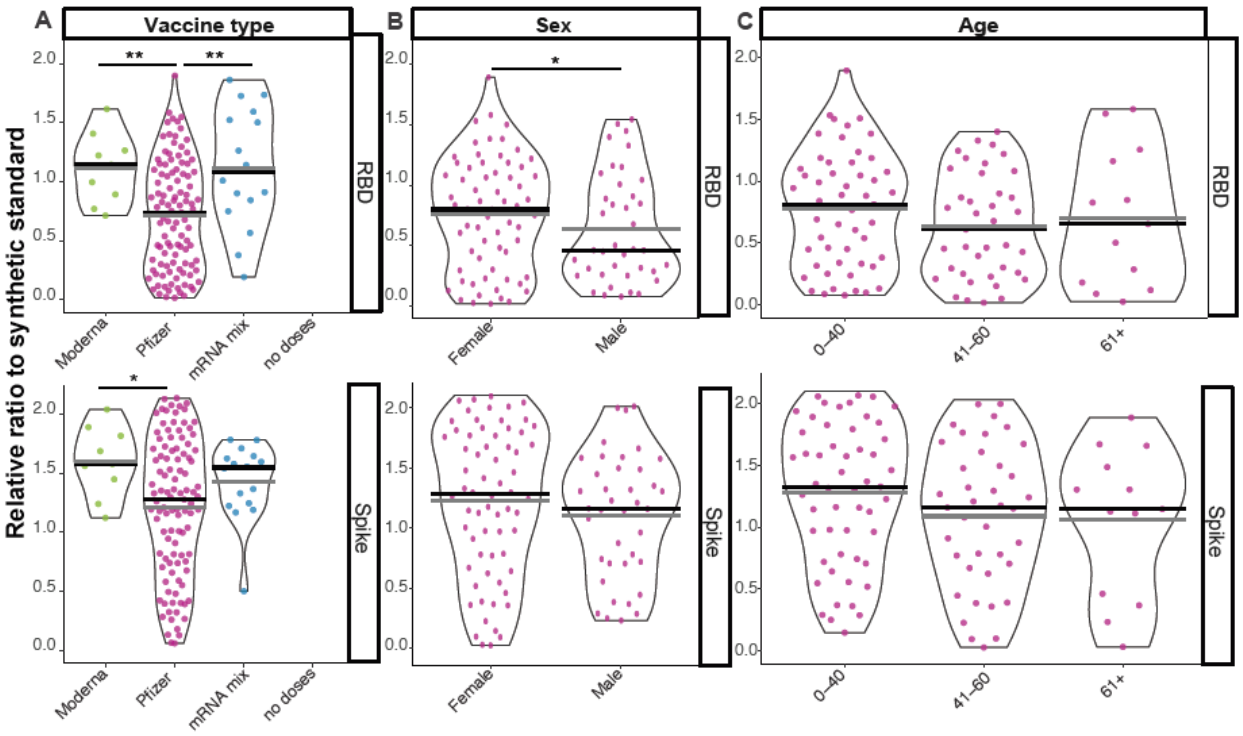


**Figure S2. Effect of variables on antibody responses.** Levels of anti-RBD and anti-spike IgG stratified by **(A)** vaccine type, (**B**) sex and (**C**) age using 0.0039 µl of sample at T3 (see figure 1A). Black lines indicate the median and gray lines signify the mean ratio value for each violin. Statistical analysis for A was for the entire cohrt, B and C for BNT162b/BNT162b group only. Unpaired comparisons were made by nonparametric Mann-Whitney test or Dunn's multiple comparisons test. **p≤0.05, **p≤0.01.*

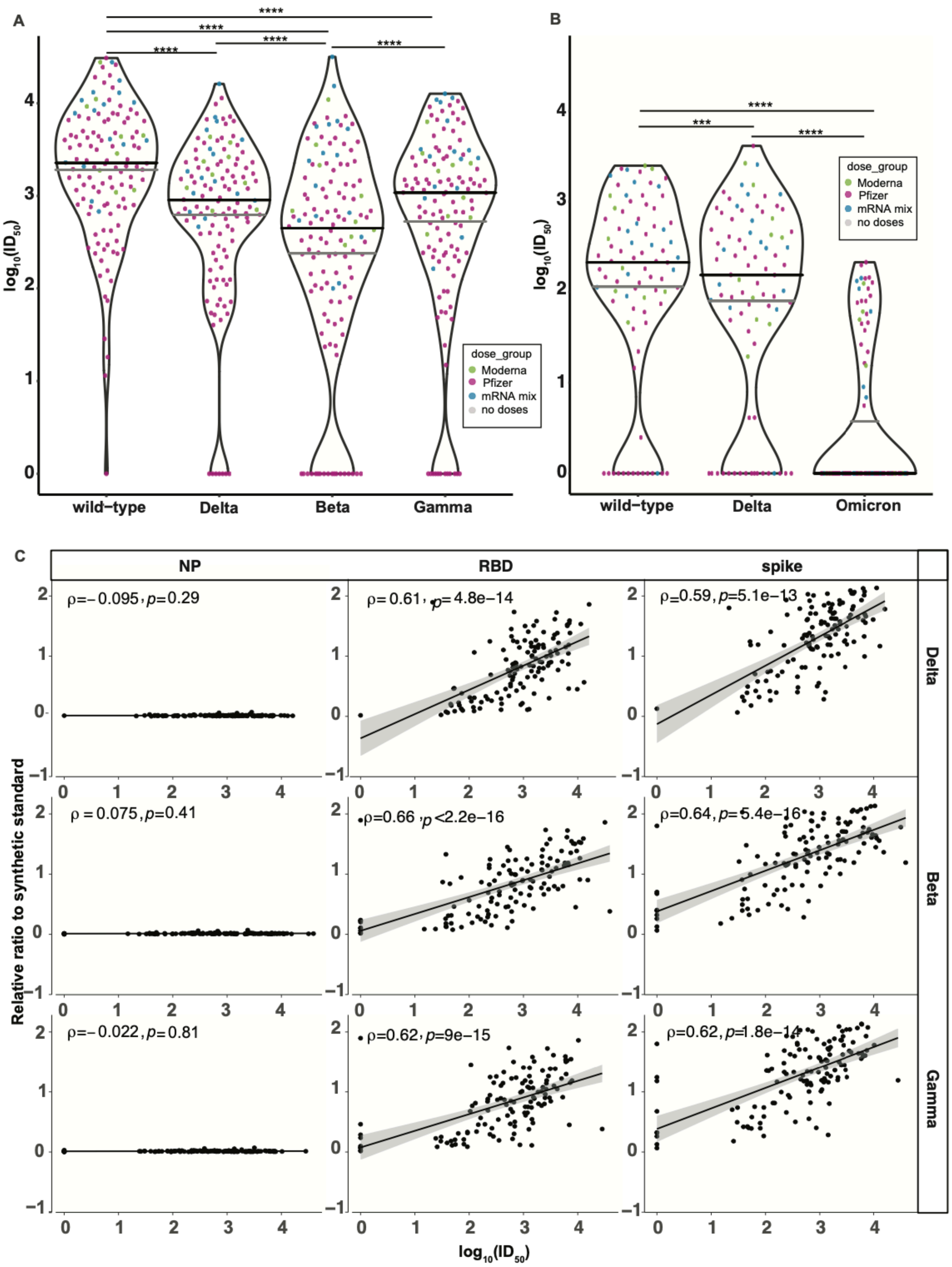
A
B
C
log10(ID50)
wild−type
Delta
Beta
Gamma
Omicron
dose_group
Moderna
Pfizer
mRNA mix
no doses
NP
RBD
spike
Relative ratio to synthetic standard
ρ=−0.095, p=0.29
ρ=0.61, p=4.8e−14
ρ=0.59, p=5.1e−13
ρ=0.075, p=0.41
ρ=0.66, p<2.2e−16
ρ=0.64, p=5.4e−16
ρ=−0.022, p=0.81
ρ=0.62, p=9e−15
ρ=0.62, p=1.8e−14

**Figure S3.** Dynamics of neutralization response of the study groups across variants after second vaccine dose. (A) T3, n=135, (B) T4, n=86. Lentiviral particles used: wildtype, B.1.617.2 (Delta), B.1.351 (Beta), P.1 (Gamma), and B. 1.1.529 (Omicron). The vaccine type is shown as colored dots (see suppl. fig. S1). Black lines indicate median and gray lines denote mean ratio value for each violin. Pairwise comparisons were made by mixed-effects ANOVA with the Geisser-Greenhouse correction, *** $p \leq 0.001$, ****$p \leq 0.0001$. **(C)** Correlation between the VOCs neutralization responses and IgG levels using the $\log_{10}$ $ID_{50}$ values of the VOCs and the relative ratio values of the spike and RBD IgG levels at T3. Solid black line is the linear regression and gray shading indicates the 95% confidence interval. P-values and Spearman's rho coefficients are indicated in each graph for the entire cohort.

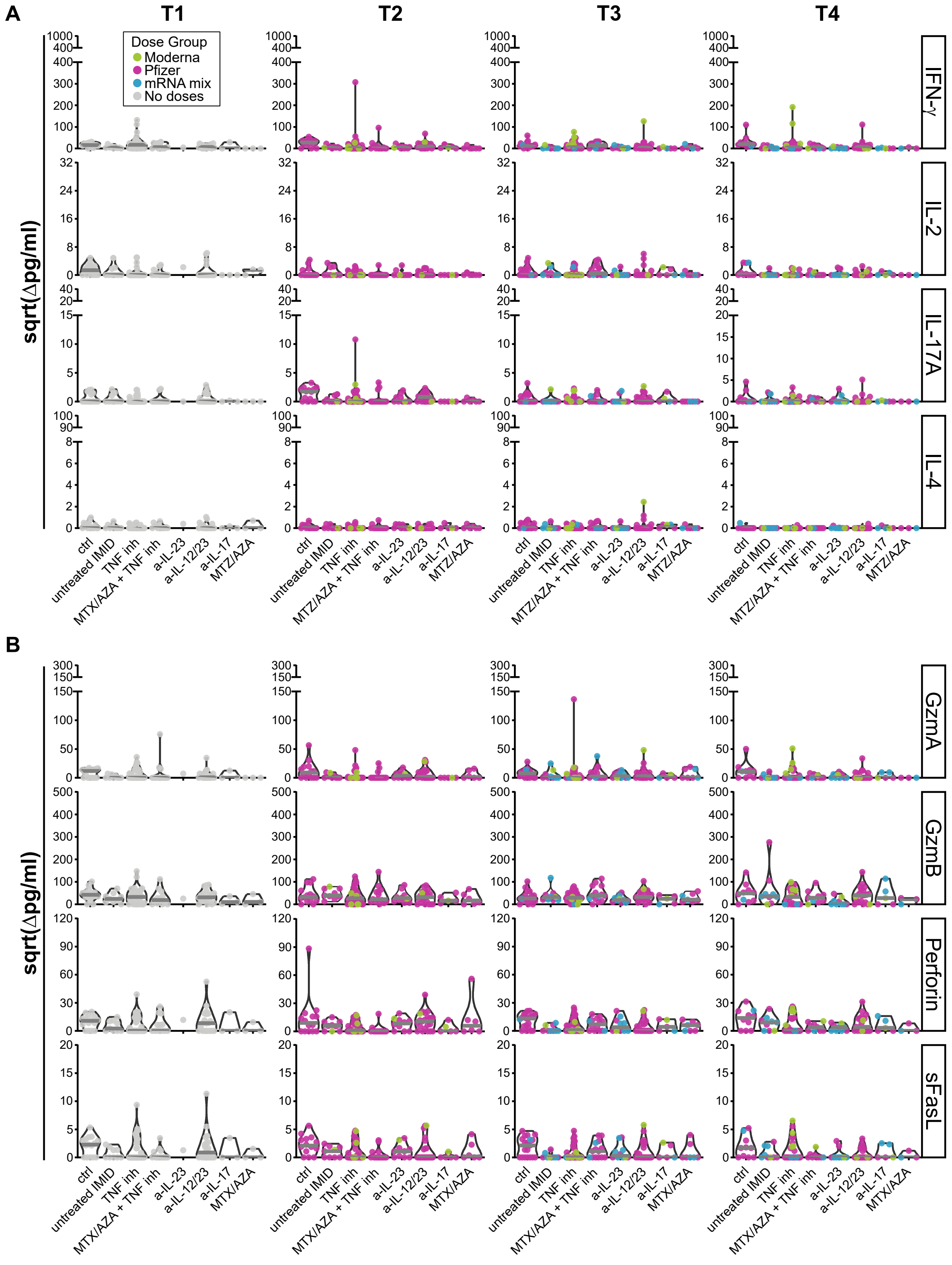
A
T1
T2
T3
T4
Dose Group
Moderna
Pfizer
mRNA mix
No doses
sqrt(Δpg/ml)
IFN-γ
IL-2
IL-17A
IL-4
ctrl
untreated IMID
TNF inh
MTX/AZA + TNF inh
MTZ/AZA + TNF inh
a-IL-23
a-IL-12/23
a-IL-17
MTZ/AZA
B
sqrt(Δpg/ml)
GzmA
GzmB
Perforin
sFasL
ctrl
untreated IMID
TNF inh
MTX/AZA + TNF inh
a-IL-23
a-IL-12/23
a-IL-17
MTX/AZA

**Figure S4.** Cellular immune responses in IMID patients to NP before or after first and second doses of mRNA vaccine. The release of cytokines and cytotoxic molecules in cell culture supernatants were analyzed by multiplex bead array following 48h stimulation with SARS-CoV-2 NP peptide pools. Violin plots show **(A)** release of cytokines IFN-γ, IL-2, IL-17A and IL-4 and **(B)** release of cytotoxic molecules GzmA, GzmB, perforin and sFasL at T1, n=102; T2, n=117; T3, n=126; and T4, n= 88, with T1 to T4 defined in figure 1A. The grey line indicates the median. Values are reported in pg/ml after subtracting background signal from wells containing PBMCs cultured with DMSO alone, as indicated by "Δ". Ctrl = Healthy controls, inh = inhibitor, MTX = methotrexate, AZA = azathioprine.

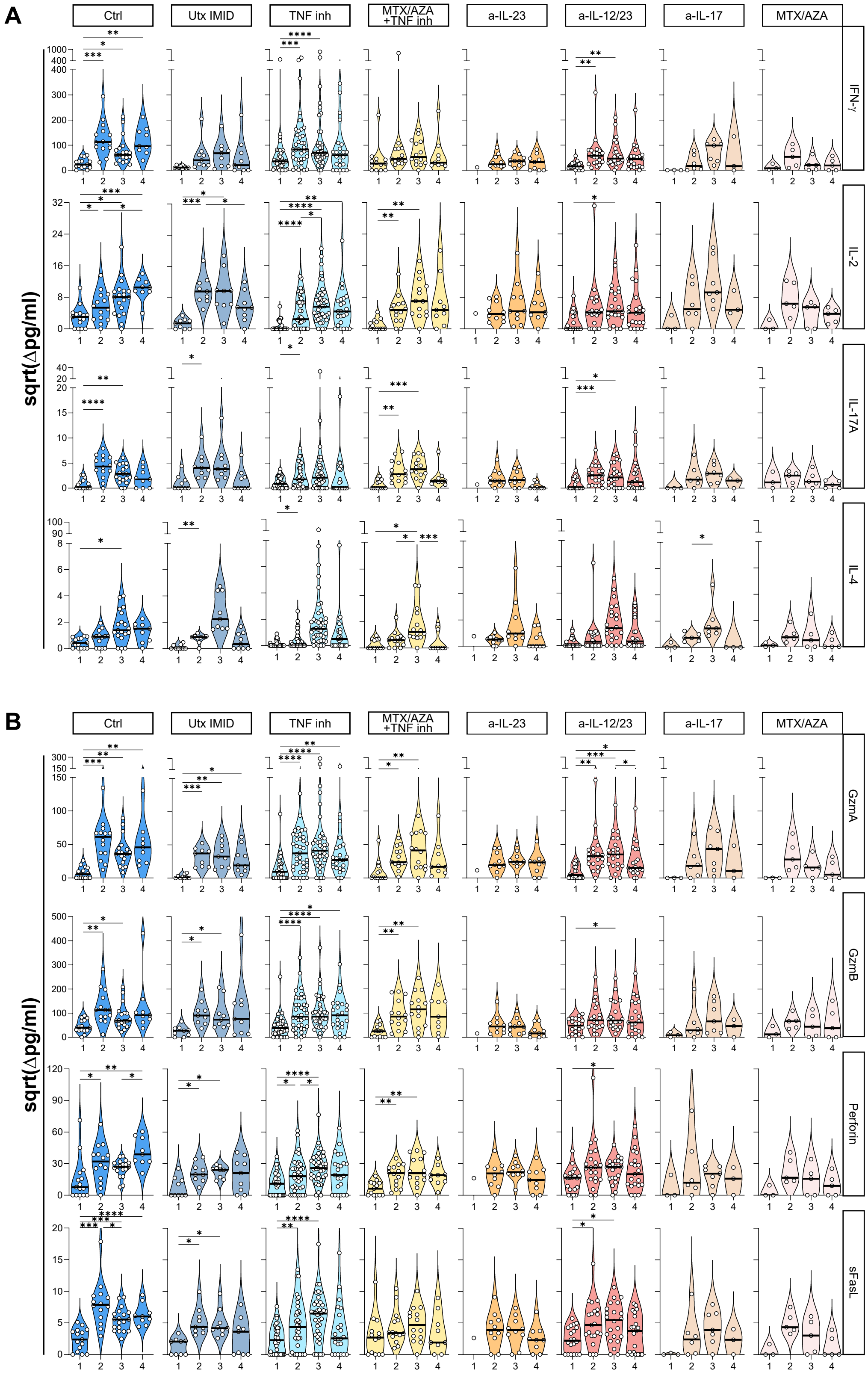
A
B
Ctrl
Utx IMID
TNF inh
MTX/AZA +TNF inh
a-IL-23
a-IL-12/23
a-IL-17
MTX/AZA
IFN-γ
IL-2
IL-17A
IL-4
GzmA
GzmB
Perforin
sFasL
sqrt(Δpg/ml)

**Figure S5.** Cytokine and cytotoxic responses in IMID patients to spike peptide pools over time. The release of cytokines and cytotoxic molecules in cell culture supernatants was analyzed by multiplex bead array following 48h stimulation with SARS-CoV-2 S peptide pools. Violin plots show release of (**A**) cytokines IFN-γ, IL-2, IL-17A and IL-4 and (**B**) cytotoxic molecules GzmA, GzmB, Perforin and sFasL at timepoints T1-T4 (see figure 1A) within each study group. The median value is indicated by the black line. Pairwise comparisons were made by mixed-effects ANOVA with the Geisser-Greenhouse correction. *$p \leq 0.05$, **$p \leq 0.01$, ***$p \leq 0.001$, ****$p \leq 0.0001$*

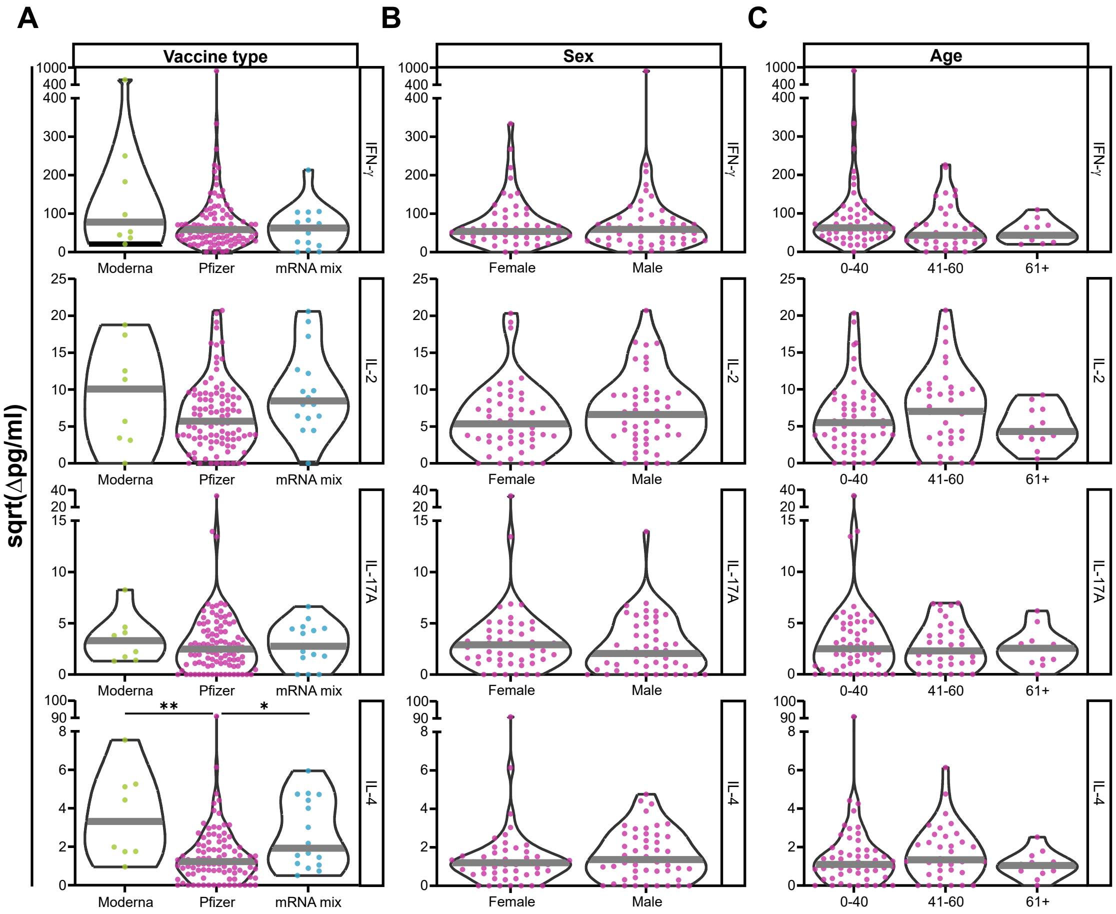


**Figure S6.** Effect of mRNA vaccine types on T cell cytokine responses to spike peptide pools in IMID patients after two doses of the indicated vaccine type. Cytokine responses in the BNT162b (Pfizer) vaccinated population were further stratified by (**A**) vaccine type (**B**) sex and (**C**) age at time point 3. Median value is indicated by the grey line in T cell cytokine violin plots. Unpaired comparisons were made by nonparametric Mann-Whitney test or Dunn's multiple comparisons test. **p≤0.05, **p≤0.01.*
